# Depression, diabetes, their comorbidity and all-cause and cause-specific mortality: a prospective cohort study

**DOI:** 10.1101/2021.12.31.21268568

**Authors:** Regina Prigge, Sarah H. Wild, Caroline A. Jackson

## Abstract

**Aims/hypothesis:** To investigate the risk of all-cause and cause-specific mortality among participants with neither, one, or both of diabetes and depression in a large prospective cohort study in the United Kingdom.

**Methods:** Our study population included 499,830 UK Biobank participants without schizophrenia and bipolar disorder at baseline. Type 1 or type 2 diabetes and depression were identified using self-reported diagnoses, prescribed medication and hospital records. Mortality was identified from death records using the primary cause of death to define cause-specific mortality. We performed Cox proportional hazards models to estimate the risk of all-cause mortality and mortality due to cancer, circulatory disease and causes of death other than circulatory disease or cancer among participants with either depression (n=41,791) or diabetes alone (n=22,677) and with comorbid diabetes and depression (n=3,597), compared to the group with neither condition (n=431,765) adjusting for sociodemographic and lifestyle factors, comorbidities, and history of CVD or cancer. We investigated for interaction between diabetes and depression.

**Results:** During a median of 6.8 (IQR: 6.1 – 7.5) years of follow-up, there were 13,724 deaths (cancer (n=7,976), circulatory disease (n=2,827), and other causes (n=2,921)). Adjusted hazard ratios of all-cause mortality and mortality due to cancer, circulatory disease and other causes were highest among people with comorbid depression and diabetes (HRs 2.16, 95% CI 1.94 – 2.42; 1.62, 95% CI 1.35 – 1.93; 2.22 95% CI 1.80 – 2.73 and 3.60, 95% CI 2.93 – 4.42, respectively). Among those with comorbid diabetes and depression, the risks of all-cause, cancer and other mortality exceeded the sum of the risks due to diabetes and depression alone.

**Conclusions/interpretation:** We confirmed the negative impact of depression and diabetes on mortality outcomes, and also identified that comorbid depression and diabetes had synergistic effects on all-cause mortality which was largely driven by deaths due to cancer and causes other than circulatory disease and cancer.

**RESEARCH IN CONTEXT:** 

**What is already known about this subject?:**

- Comorbid depression is common in individuals with diabetes and associated with increased risks of all-cause and cardiovascular mortality
- The mortality risk among people with comorbid diabetes and depression might exceed the sum of the risks associated with each disorder alone
- There is limited knowledge about the individual and joint effects of depression and diabetes on risk of death from specific causes.

**What is the key question?:**

- What is the risk of all-cause and cause-specific mortality associated with neither, one, or both of diabetes (of any type) and depression?

**What are the new findings?:**

- In a large prospective cohort study in the United Kingdom, comorbid depression and diabetes had synergistic effects on all-cause mortality which was largely driven by deaths due to cancer and causes other than circulatory disease and cancer

**How might this impact on clinical practice in the foreseeable future?:**

- These findings help identify individuals at high risk of adverse events, and suggest a need for cost-effective interventions to support psychological well-being and risk reduction in people with diabetes

## INTRODUCTION

Depression is common among individuals with diabetes with about 34% of women and 23% of men with type 2 diabetes having comorbid depression [1]. As such, individuals with diabetes are disproportionally affected by depression compared to the general population [1-3]. Importantly, those affected by both conditions are at higher risk of poor glycaemic control [4], medical non-compliance [5, 6], and at greater risks of micro-and macrovascular complications [7, 8], relative to those with diabetes alone.

Previous meta-analyses have shown that comorbid depression is associated with approximately 50 – 75% increased risk of all-cause mortality and 50% increased risk of cardiovascular mortality in individuals with type 1 or type 2 diabetes [9-13]. Importantly, since all of the included studies were based on patients with diabetes, they could not assess whether the presence of diabetes modified the excess mortality associated with depression.

There is some evidence that the association between depression and all-cause mortality may be more pronounced among individuals with versus without type 1 or type 2 diabetes [8, 14, 15] but few existing studies have investigated individual and joint effects of depression and diabetes on mortality risk [7, 14, 16-20]. These studies consistently reported that people with comorbid depression and diabetes were at particularly high risk of all-cause and cardiac mortality. Furthermore, they largely suggested synergistic effects (supra-additive interaction) between depression and diabetes on risk of all-cause mortality and, to a lesser extent, on risk of cardiac mortality. None of the existing studies in the general population have investigated causes of death other than circulatory disease, despite evidence that depression may be linked to an increased risk of non-cardiovascular, non-cancer mortality in patients with diabetes [21]. Furthermore, existing studies are largely based on samples from the United States [7, 17-20], are limited by small sample sizes [7, 16, 17] or are based on a selected patient population, such as patients who were hospitalised for myocardial infarction [16, 19].

Despite the substantial burden of both depression and diabetes, and the potential impact on the prognosis of patients affected by both disorders, there is limited knowledge about the individual and joint effects of depression and diabetes on risk of death from specific causes. Therefore, the aim of this study was to investigate the risk of all-cause and cause-specific mortality among participants with neither, one or both of diabetes and depression in a large prospective cohort in the United Kingdom.

## RESEARCH DESIGN AND METHODS

### Study population

We included participants from the UK Biobank, a prospective cohort study of ∼500,000 participants aged 40 – 69 years at baseline, recruited in 22 assessment centres in England, Scotland, and Wales from 2006 to 2010 and followed up through linkage to routinely available national datasets [22]. Cohort entry date forms the baseline for this study. We excluded participants who withdrew from the study, whose information could not be linked to hospital or death records and those with a history of bipolar disorder or schizophrenia at baseline. Analyses of UK Biobank data are conducted under generic approval from the NHS National Research Ethics Service (Ref 11/NW/0382, approval letter dated 17 June 2011). Full written informed consent was obtained from participants at the point of data collection.

### Exposure: Depression, diabetes, and their comorbidity

We defined depression as at least one of: self-reported antidepressant use; self-reported depression; or hospital record of depression at baseline. Antidepressant use and self-reported doctor-diagnosis of depression were identified in the nurse interview at baseline. In line with previous algorithms, we defined antidepressant use as self-reported use of at least one selective serotonin reuptake inhibitor or other antidepressant medication [23]. We defined hospital record of depression as diagnosis with depressive episode or recurrent depressive episode in hospital records (ICD 10: F32.X, F33.X) prior to recruitment to the study and diabetes as self-reported diagnosis with, and/or treatment for, type 1 or 2 diabetes, or hospital record of type 1 or 2 diabetes at baseline (ICD 10: E10.X – E14.X). Self-reported diabetes was ascertained in the touchscreen questionnaire and nurse interview. Anti-diabetic treatment was defined as self-report of any medication listed in ‘A10 Drugs used in diabetes’ of the ATC/DDD Index 2018 [24]. We used information on depression and diabetes at baseline to create our exposure variable consisting of four levels: neither depression nor diabetes, depression alone, diabetes alone, and both depression and diabetes.

### Outcome: All-cause and cause-specific mortality

We ascertained mortality from linked death records using the primary or underlying cause of death to define cause-specific mortality. Causes and dates of death were provided by NHS Digital for participants from England and Wales, and by the Information and Statistics Division for participants from Scotland [25]. We defined circulatory deaths using ICD 10 codes I00 – I99 and G.45, cancer deaths using ICD 10 codes C00 – C97 and other mortality as deaths due to any other cause. We calculated survival times from the date of attending the baseline assessment centre to the date of death, or end of follow-up (31 November 2015).

### Covariates

Information on sociodemographic and lifestyle factors, comorbidities, and family history were ascertained through self-report in the touchscreen questionnaire and nurse interview and, where available, through measured values at the baseline assessment centre (ESM Material S1). Covariates included age, sex, ethnicity, income, education, area-based deprivation, body mass index (BMI), smoking, alcohol intake, physical activity, fruit and vegetable intake, hypertension, high cholesterol, history of cardiovascular disease (CVD), history of cancer, family history of CVD, and family history of severe depression.

### Statistical analyses

We performed analyses using R version 3.6.2. We assessed baseline characteristics across the four exposure groups. Due to the large sample size, we did not perform formal tests for differences between groups. We used Cox proportional hazards models to estimate hazard ratios and 95% confidence intervals (CI) of the risk of all-cause and cause-specific mortality among participants with either diabetes or depression alone and with comorbid diabetes and depression, relative to the group with neither condition. The first model describes the unadjusted association, the second is adjusted for age, sex, ethnicity, and socioeconomic factors (education, income, and area-based deprivation), and the third additionally controls for BMI, alcohol intake, physical activity, smoking, fruit and vegetable consumption, oily fish intake, family history of CVD or depression, and history of hypertension, high cholesterol levels, history of CVD and cancer at baseline. We performed pre-specified sex-stratified analyses to evaluate differences between men and women. Since age did not fulfil the assumption of a linear contribution to the Cox proportional hazards model, we split the age distribution into four equally sized groups and included age as a categorical variable in the models. We tested the proportional hazards assumption for all variables using the cox.zph function, and investigated potential violations using log-minus-log survival plots and plots of scaled Schoenfeld residuals against time. We allowed for different baseline hazards for covariates that did not meet the proportional hazards assumption. Nonetheless, there was evidence of a violation of the proportional hazards assumption due to small numbers of events towards the end of follow-up in one analysis. Since truncation of follow-up at six years gave similar point estimates, the results for the whole follow-up are presented (ESM Table S1).

We tested for multiplicative interaction by adding a product term of depression and diabetes to the fully adjusted Cox proportional hazards model (and considered a two-sided p<0.05 statistically significant). We also investigated for additive (i.e. biological) interaction, which is more important for understanding population health, by calculating the relative excess risk due to interaction (RERI), the attributable proportion due to interaction (AP), and the synergy index (S), with accompanying 95% CIs [26].

Differences between participants with and without complete data were indicative of a violation of the missing completely at random (MCAR) assumption (ESM Table S2). Whilst the average number of variables with missing information among participants was low, overall missingness was high due to the large number of included variables, with 155,761 (31.2%) participants having at least one missing value in any variable. Since a missing at random mechanism was deemed likely, multiple imputation of missing data was performed using the MICE package in R [27]. In keeping with the recommendation that the number of imputations should equate to the percent of incomplete cases [28], we performed 32 imputations with 10 iterations. The imputations were run separately for participants with and without depression to take account of our interest in the interaction between depression and diabetes. Since a complete case analysis is likely to be biased when missing data are not MCAR, the primary analyses are based on imputed data, with results of the complete case analysis provided in ESM Tables S3 – S6.

### Sensitivity analysis

To facilitate comparisons with previous studies, we also performed a sensitivity analysis with sub-groups of circulatory mortality as the outcome, specifically CVD and non-CVD circulatory mortality. CVD mortality was defined as death due to ischaemic heart disease, cerebrovascular disease, or transient ischaemic attack (ICD 10: I20.X – I25.X, I60.X – I69.X, G45).

## RESULTS

### Descriptive statistics

After excluding people with schizophrenia and bipolar disorder, our study population included 499,830 people (Fig. 1), with a median age of 58 (IQR: 50 – 63) years at cohort entry, of whom 227,794 (45.6%) were male, and 470,282 (94.1%) had a white ethnic background. At baseline, 431,765 (86.4%) participants had neither depression nor diabetes, 41,791 (8.4%) had depression alone, 22,677 (4.5%) had diabetes alone, and 3,597 (0.7%) had both diabetes and depression.

**Fig. 1.**
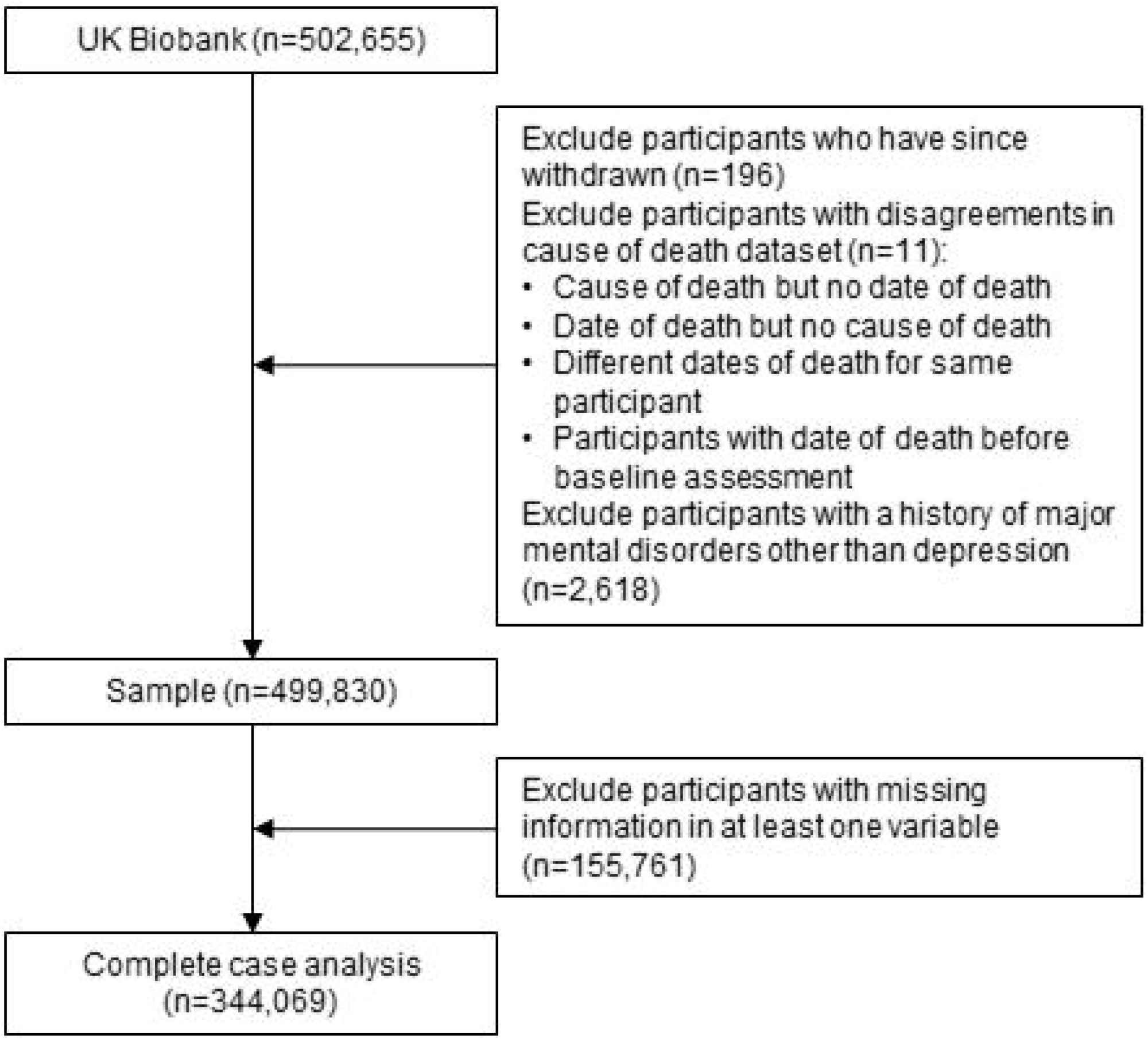
Flow diagram of sample selection

Baseline characteristics differed between exposure groups (Table 1). The group with diabetes alone was older and included a higher proportion of men than other groups. Compared to the other groups, the group with comorbid depression and diabetes had the highest proportion with low socioeconomic status and higher prevalence of cardiovascular risk factors in general.

**Table 1:**
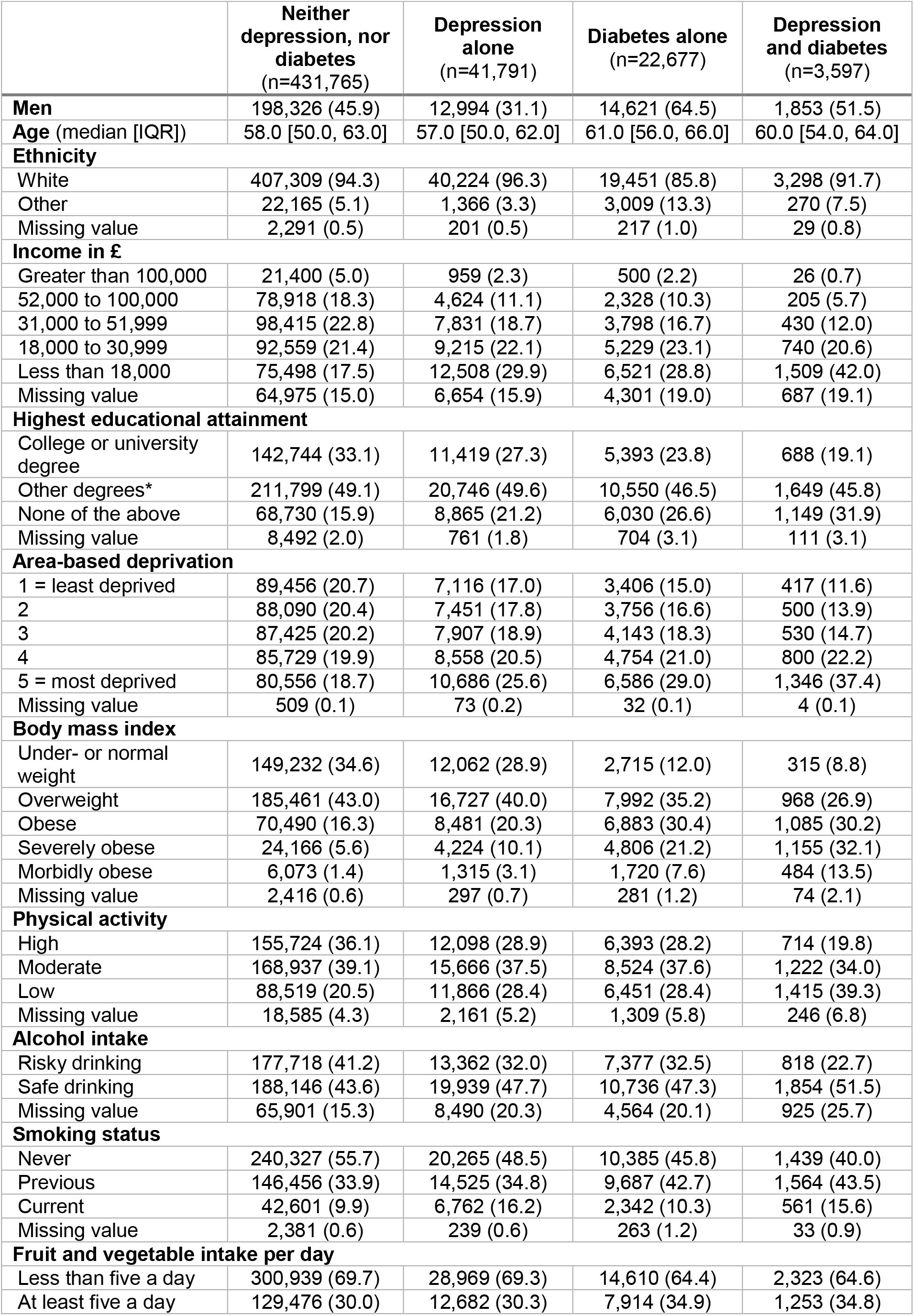
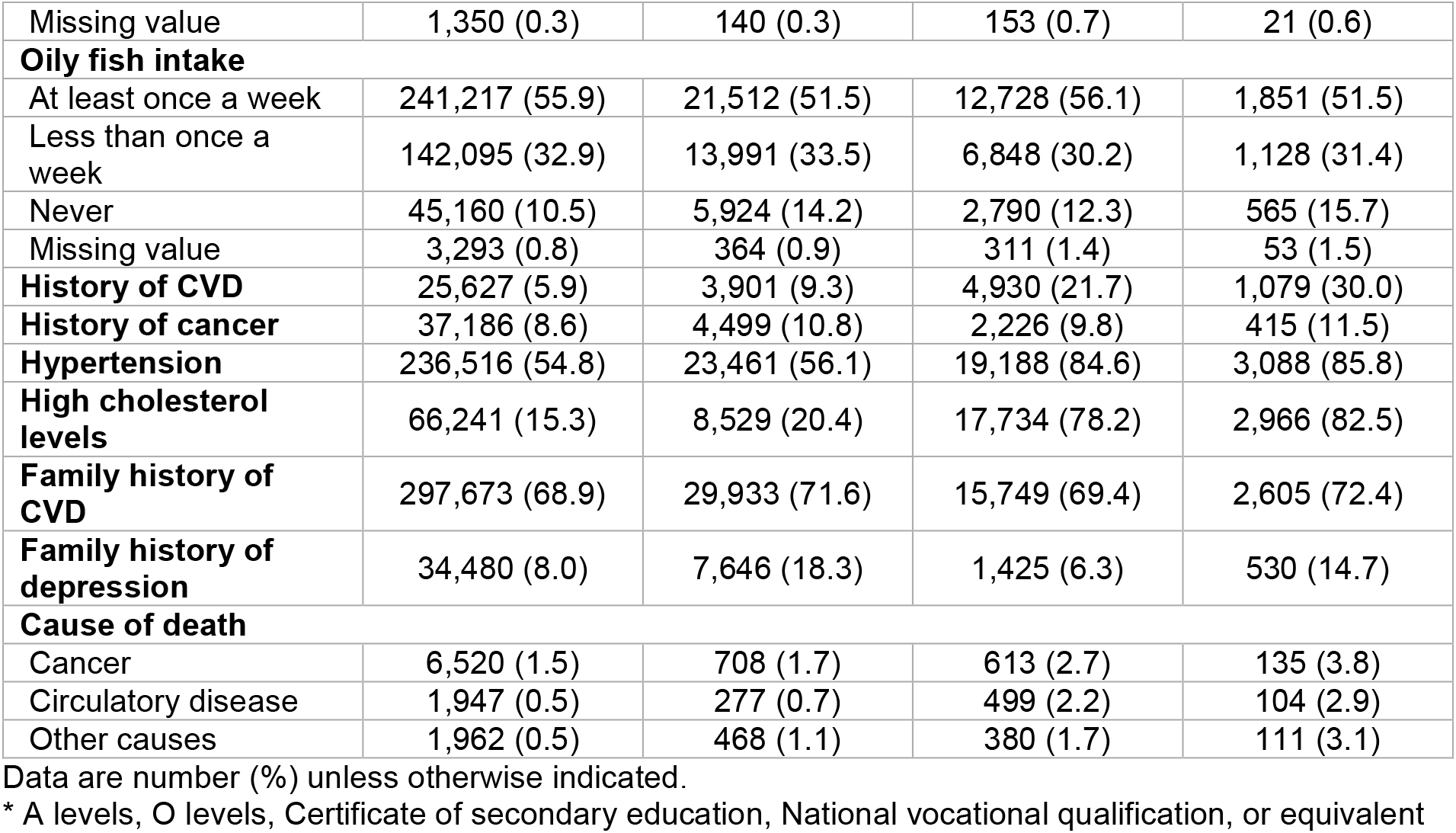
Baseline characteristics and causes of death for UK Biobank participants with neither, one or both of depression and diabetes

|  | **Neither**<br>**depression, nor**<br>**diabetes**<br>**(n=431,765)** | **Depression**<br>**alone**<br>**(n=41,791)** | **Diabetes alone**<br>**(n=22,677)** | **Depression**<br>**and diabetes**<br>**(n=3,597)** |
| --- | --- | --- | --- | --- |
| **Men** | 198,326 (45.9) | 12,994 (31.1) | 14,621 (64.5) | 1,853 (51.5) |
| **Age** (median [IQR]) | 58.0 [50.0, 63.0] | 57.0 [50.0, 62.0] | 61.0 [56.0, 66.0] | 60.0 [54.0, 64.0] |
| **Ethnicity** |  |  |  |  |
| White | 407,309 (94.3) | 40,224 (96.3) | 19,451 (85.8) | 3,298 (91.7) |
| Other | 22,165 (5.1) | 1,366 (3.3) | 3,009 (13.3) | 270 (7.5) |
| Missing value | 2,291 (0.5) | 201 (0.5) | 217 (1.0) | 29 (0.8) |
| **Income in £** |  |  |  |  |
| Greater than 100,000 | 21,400 (5.0) | 959 (2.3) | 500 (2.2) | 26 (0.7) |
| 52,000 to 100,000 | 78,918 (18.3) | 4,624 (11.1) | 2,328 (10.3) | 205 (5.7) |
| 31,000 to 51,999 | 98,415 (22.8) | 7,831 (18.7) | 3,798 (16.7) | 430 (12.0) |
| 18,000 to 30,999 | 92,559 (21.4) | 9,215 (22.1) | 5,229 (23.1) | 740 (20.6) |
| Less than 18,000 | 75,498 (17.5) | 12,508 (29.9) | 6,521 (28.8) | 1,509 (42.0) |
| Missing value | 64,975 (15.0) | 6,654 (15.9) | 4,301 (19.0) | 687 (19.1) |
| **Highest educational attainment** |  |  |  |  |
| College or university degree | 142,744 (33.1) | 11,419 (27.3) | 5,393 (23.8) | 688 (19.1) |
| Other degrees* | 211,799 (49.1) | 20,746 (49.6) | 10,550 (46.5) | 1,649 (45.8) |
| None of the above | 68,730 (15.9) | 8,865 (21.2) | 6,030 (26.6) | 1,149 (31.9) |
| Missing value | 8,492 (2.0) | 761 (1.8) | 704 (3.1) | 111 (3.1) |
| **Area-based deprivation** |  |  |  |  |
| 1 = least deprived | 89,456 (20.7) | 7,116 (17.0) | 3,406 (15.0) | 417 (11.6) |
| 2 | 88,090 (20.4) | 7,451 (17.8) | 3,756 (16.6) | 500 (13.9) |
| 3 | 87,425 (20.2) | 7,907 (18.9) | 4,143 (18.3) | 530 (14.7) |
| 4 | 85,729 (19.9) | 8,558 (20.5) | 4,754 (21.0) | 800 (22.2) |
| 5 = most deprived | 80,556 (18.7) | 10,686 (25.6) | 6,586 (29.0) | 1,346 (37.4) |
| Missing value | 509 (0.1) | 73 (0.2) | 32 (0.1) | 4 (0.1) |
| **Body mass index** |  |  |  |  |
| Under- or normal weight | 149,232 (34.6) | 12,062 (28.9) | 2,715 (12.0) | 315 (8.8) |
| Overweight | 185,461 (43.0) | 16,727 (40.0) | 7,992 (35.2) | 968 (26.9) |
| Obese | 70,490 (16.3) | 8,481 (20.3) | 6,883 (30.4) | 1,085 (30.2) |
| Severely obese | 24,166 (5.6) | 4,224 (10.1) | 4,806 (21.2) | 1,155 (32.1) |
| Morbidly obese | 6,073 (1.4) | 1,315 (3.1) | 1,720 (7.6) | 484 (13.5) |
| Missing value | 2,416 (0.6) | 297 (0.7) | 281 (1.2) | 74 (2.1) |
| **Physical activity** |  |  |  |  |
| High | 155,724 (36.1) | 12,098 (28.9) | 6,393 (28.2) | 714 (19.8) |
| Moderate | 168,937 (39.1) | 15,666 (37.5) | 8,524 (37.6) | 1,222 (34.0) |
| Low | 88,519 (20.5) | 11,866 (28.4) | 6,451 (28.4) | 1,415 (39.3) |
| Missing value | 18,585 (4.3) | 2,161 (5.2) | 1,309 (5.8) | 246 (6.8) |
| **Alcohol intake** |  |  |  |  |
| Risky drinking | 177,718 (41.2) | 13,362 (32.0) | 7,377 (32.5) | 818 (22.7) |
| Safe drinking | 188,146 (43.6) | 19,939 (47.7) | 10,736 (47.3) | 1,854 (51.5) |
| Missing value | 65,901 (15.3) | 8,490 (20.3) | 4,564 (20.1) | 925 (25.7) |
| **Smoking status** |  |  |  |  |
| Never | 240,327 (55.7) | 20,265 (48.5) | 10,385 (45.8) | 1,439 (40.0) |
| Previous | 146,456 (33.9) | 14,525 (34.8) | 9,687 (42.7) | 1,564 (43.5) |
| Current | 42,601 (9.9) | 6,762 (16.2) | 2,342 (10.3) | 561 (15.6) |
| Missing value | 2,381 (0.6) | 239 (0.6) | 263 (1.2) | 33 (0.9) |
| **Fruit and vegetable intake per day** |  |  |  |  |
| Less than five a day | 300,939 (69.7) | 28,969 (69.3) | 14,610 (64.4) | 2,323 (64.6) |
| At least five a day | 129,476 (30.0) | 12,682 (30.3) | 7,914 (34.9) | 1,253 (34.8) |
| Missing value | 1,350 (0.3) | 140 (0.3) | 153 (0.7) | 21 (0.6) |
| **Oily fish intake** |  |  |  |  |
| At least once a week | 241,217 (55.9) | 21,512 (51.5) | 12,728 (56.1) | 1,851 (51.5) |
| Less than once a week | 142,095 (32.9) | 13,991 (33.5) | 6,848 (30.2) | 1,128 (31.4) |
| Never | 45,160 (10.5) | 5,924 (14.2) | 2,790 (12.3) | 565 (15.7) |
| Missing value | 3,293 (0.8) | 364 (0.9) | 311 (1.4) | 53 (1.5) |
| **History of CVD** | 25,627 (5.9) | 3,901 (9.3) | 4,930 (21.7) | 1,079 (30.0) |
| **History of cancer** | 37,186 (8.6) | 4,499 (10.8) | 2,226 (9.8) | 415 (11.5) |
| **Hypertension** | 236,516 (54.8) | 23,461 (56.1) | 19,188 (84.6) | 3,088 (85.8) |
| **High cholesterol levels** | 66,241 (15.3) | 8,529 (20.4) | 17,734 (78.2) | 2,966 (82.5) |
| **Family history of CVD** | 297,673 (68.9) | 29,933 (71.6) | 15,749 (69.4) | 2,605 (72.4) |
| **Family history of depression** | 34,480 (8.0) | 7,646 (18.3) | 1,425 (6.3) | 530 (14.7) |
| **Cause of death** |  |  |  |  |
| Cancer | 6,520 (1.5) | 708 (1.7) | 613 (2.7) | 135 (3.8) |
| Circulatory disease | 1,947 (0.5) | 277 (0.7) | 499 (2.2) | 104 (2.9) |
| Other causes | 1,962 (0.5) | 468 (1.1) | 380 (1.7) | 111 (3.1) |

Data are number (%) unless otherwise indicated.
\* A levels, O levels, Certificate of secondary education, National vocational qualification, or equivalent

During a median of 6.8 (IQR 6.1 – 7.5) years of follow-up, there were 13,724 deaths, 7,976 of which were due to cancer, and 2,827 and 2,921 due to circulatory disease and other causes, respectively. Overall, the three most frequent other causes of death were respiratory diseases, diseases of the digestive system and external causes of mortality. A detailed description of other causes of death by exposure group is presented in ESM Table S7. For all causes of deaths, crude proportions of participants who died were lowest in the group with neither depression nor diabetes, followed by the group with depression alone and diabetes alone and highest among those with both depression and diabetes (Table 1).

### Associations with all-cause and cause-specific mortality

Diabetes alone and comorbid diabetes and depression were associated with greater risks of all-cause and all cause-specific mortality relative to participants with neither depression nor diabetes in all models, whereas depression alone was not associated with greater risks of cancer mortality (Table 2). The associations attenuated but remained statistically significant after adjusting for various factors. In the fully adjusted model, compared to those with neither condition, the risk of all-cause mortality was 26% higher among people with depression alone (HR 1.26, 95% CI 1.19 – 1.33), 62% higher among individuals with diabetes alone (HR 1.62, 95% CI 1.52 – 1.72), and 116% higher among people with comorbid diabetes and depression (HR 2.16, 95% CI 1.94 – 2.42). In all models, the associations followed the same pattern with the largest effect sizes for those with both depression and diabetes and lowest for those with depression alone.

**Table 2:** Hazard ratios (95% CI) for all-cause and cause-specific mortality among UK Biobank participants with neither, one of both of depression and diabetes

|  |  | **Unadjusted HR**<br>**(95% CI)** | **Adjusted HR**<br>**(95% CI)**<br>**Model 1\*** | **Adjusted HR**<br>**(95% CI)**<br>**Model 2†** |
| --- | --- | --- | --- | --- |
| **All-cause mortality** | Neither depression, nor diabetes | 1.0 | 1.0 | 1.0 |
|  | Depression alone | 1.44 (1.36 – 1.52) | 1.45 (1.37 – 1.53) | 1.26 (1.19 – 1.33) |
|  | Diabetes alone | 2.82 (2.67 – 2.97) | 1.86 (1.76 – 1.97) | 1.62 (1.52 – 1.72) |
|  | Depression and diabetes | 4.19 (3.77 – 4.66) | 2.88 (2.59 – 3.21) | 2.16 (1.94 – 2.42) |
| **Cancer mortality** | Neither depression, nor diabetes | 1.0 | 1.0 | 1.0 |
|  | Depression alone | 1.12 (1.04 – 1.21) | 1.12 (1.04 – 1.22) | 1.00 (0.92 – 1.08) |
|  | Diabetes alone | 1.85 (1.70 – 2.01) | 1.31 (1.21 – 1.43) | 1.24 (1.13 – 1.36) |
|  | Depression and diabetes | 2.58 (2.18 – 3.06) | 1.91 (1.61 – 2.27) | 1.62 (1.35 – 1.93) |
| **Circulatory mortality** | Neither depression, nor diabetes | 1.0 | 1.0 | 1.0 |
|  | Depression alone | 1.47 (1.30 – 1.67) | 1.56 (1.37 – 1.77) | 1.27 (1.12 – 1.45) |
|  | Diabetes alone | 5.04 (4.57 – 5.56) | 2.89 (2.61 – 3.20) | 1.97 (1.76 – 2.20) |
|  | Depression and diabetes | 6.66 (5.47 – 8.11) | 4.12 (3.37 – 5.03) | 2.22 (1.80 – 2.73) |
| **Mortality due to other causes** | Neither depression, nor diabetes | 1.0 | 1.0 | 1.0 |
|  | Depression alone | 2.46 (2.23 – 2.72) | 2.38 (2.14 – 2.63) | 2.10 (1.89 – 2.34) |
|  | Diabetes alone | 3.82 (3.43 – 4.27) | 2.41 (2.16 – 2.70) | 2.23 (1.97 – 2.53) |
|  | Depression and diabetes | 7.07 (5.84 – 8.57) | 4.43 (3.65 – 5.38) | 3.60 (2.93 – 4.42) |

\*Model 1: Age, sex, ethnicity, education, income, area-based deprivation
†Model 2: Model 1 + BMI, physical activity, alcohol intake, smoking, fruit and vegetable intake, oily fish intake, cholesterol, hypertension, cardiovascular disease, cancer, family history of cardiovascular disease, and family history of depression

The associations for the groups with depression alone and diabetes alone were mostly similar in men and women (Table 3). However, the relative risk of circulatory mortality among those with diabetes alone and the relative risk of other mortality among those with depression and diabetes were higher for women than men. However, confidence intervals were wide and overlapped.

**Table 3:** Hazard ratios (95% CI) for all-cause and cause-specific mortality among UK Biobank participants with neither, one of both of depression and diabetes, separately for men and women

|  |  | **Men**<br>Adjusted HR (95% CI)* | **Women**<br>Adjusted HR (95% CI)* |
| --- | --- | --- | --- |
| **All-cause mortality** | Neither depression, nor diabetes | 1.0 | 1.0 |
|  | Depression alone | 1.30 (1.20 – 1.41) | 1.24 (1.14 – 1.34) |
|  | Diabetes alone | 1.63 (1.52 – 1.75) | 1.59 (1.41 – 1.80) |
|  | Depression and diabetes | 2.10 (1.83 – 2.41) | 2.28 (1.88 – 2.76) |
| **Cancer mortality** | Neither depression, nor diabetes | 1.0 | 1.0 |
|  | Depression alone | 0.96 (0.85 – 1.09) | 1.04 (0.94 – 1.15) |
|  | Diabetes alone | 1.23 (1.11 – 1.38) | 1.26 (1.07 – 1.49) |
|  | Depression and diabetes | 1.63 (1.30 – 2.03) | 1.58 (1.18 – 2.12) |
| **Circulatory mortality** | Neither depression, nor diabetes | 1.0 | 1.0 |
|  | Depression alone | 1.32 (1.12 – 1.55) | 1.21 (0.97 – 1.51) |
|  | Diabetes alone | 1.91 (1.69 – 2.16) | 2.28 (1.77 – 2.95) |
|  | Depression and diabetes | 2.22 (1.74 – 2.82) | 2.24 (1.45 – 3.46) |
| **Mortality due to other causes** | Neither depression, nor diabetes | 1.0 | 1.0 |
|  | Depression alone | 2.11 (1.83 – 2.44) | 2.08 (1.78 – 2.44) |
|  | Diabetes alone | 2.27 (1.97 – 2.63) | 2.19 (1.69 – 2.83) |
|  | Depression and diabetes | 3.13 (2.40 – 4.08) | 4.51 (3.24 – 6.27) |

\*Fully adjusted model: Age, ethnicity, education, income, area-based deprivation, BMI, physical activity, alcohol intake, smoking, fruit and vegetable intake, oily fish intake, cholesterol, hypertension, cardiovascular disease, cancer, family history of cardiovascular disease, and family history of depression

### Additive and multiplicative interaction

Among the group with comorbid diabetes and depression, the risks of all-cause, cancer, and other mortality exceeded the sum of the risks due to diabetes and depression alone (ESM Fig. S1), suggesting the presence of an interaction on the additive scale. However, formal statistical evidence of additive interaction was only found for all-cause and cancer mortality (relative excess risk due to interaction: 0.29 (95% CI 0.03 – 0.54) and 0.38 (95% CI 0.07 – 0.68), respectively) (ESM Table S8). Furthermore, there was evidence of interaction between depression and diabetes on the multiplicative scale for cancer mortality (p = 0.006) but not for all-cause mortality, circulatory mortality and mortality from other causes (p = 0.182, 0.578 and 0.061, respectively).

### Sensitivity analyses

Depression alone, diabetes alone and comorbid diabetes and depression were associated with greater risks of both CVD and non-CVD circulatory mortality relative to participants with neither depression nor diabetes in unadjusted and partially adjusted models. In the fully adjusted model, the association with CVD and non-CVD circulatory mortality was strongest among the group with comorbid depression and diabetes. Whilst the association with CVD mortality was weakest among those with depression alone, the association with non-CVD circulatory mortality was similar for those with depression alone and diabetes alone. For non-CVD circulatory mortality, the risk among the group with comorbid depression and diabetes exceeded the sum of the risks due to depression and diabetes alone (ESM Table S9). There was no evidence for multiplicative or additive interaction for either CVD or non-CVD circulatory mortality (ESM Table S8).

## DISCUSSION

### Principal findings

In a large prospective study in the United Kingdom, we confirmed the higher mortality associated with each of depression and diabetes, and also identified synergistic effects of depression and diabetes on different causes of mortality beyond that expected from their individual effects. This pattern remained even after adjusting for a wide range of confounding factors.

### Strengths and limitations of this study

Our study has a number of strengths. It is one of just a few cohort studies investigating the relative importance and synergistic effects of depression and diabetes on all-cause and circulatory mortality in the general population and within a universal healthcare setting. Furthermore, to our knowledge, this is the first study to describe the effects of comorbid depression and diabetes on mortality from cancer and causes of death other than circulatory disease or cancer. We used a large prospective cohort that contained detailed information on a range of potential confounding factors. The large sample size and subsequent large number of deaths meant we had sufficient power to investigate the individual and synergistic effects of our exposure groups, different causes of deaths and stratify our analysis by sex. A further advantage of our study is that, in contrast to many other prospective cohort studies, there was limited attrition, because we relied on administrative health records to ascertain outcomes.

Our study has some limitations. The UK Biobank had a low response rate (5.5%) which resulted in a relatively healthy cohort from a higher socioeconomic background than the general population [32]. However, it has been argued that this is unlikely to influence estimates of associations between diseases given that there are large numbers of participants with different levels of risk factors in the sample [32]. Nonetheless, selection bias might have influenced some results of this analysis. Also, there is potential for misclassification because participants might have underreported depression, diabetes and their comorbidity at baseline. Although our measurement at baseline used hospital records as well as self-report, it is possible that we misclassified some participants’ exposure status and have underestimated the mortality risks associated with depression and diabetes. We were not able to update exposure status during follow-up. Also, we may have missed a small number of deaths occurring outside the UK, but this is likely to have occurred non-differentially across the four exposure groups. This may however have further biased our findings towards the null. Although key confounding factors were adjusted for in this analysis, residual confounding might explain some of the observed effect, for example if the measurement error of lifestyle factors and comorbidities was systematically different among the four exposure groups.

### Strengths and weaknesses in relation to other studies

Our findings are in keeping with previous studies reporting a high risk of all-cause and circulatory mortality among people with comorbid depression and diabetes that exceeds the risk due to having either diabetes or depression alone [7, 14, 16-20]. Whilst the strength of the association between comorbid depression and diabetes and all-cause and circulatory mortality was similar in some previous studies [16-18, 20], others observed much higher HRs of 3.64 [19], 3.71 [14], and 4.56 [7] for all-cause mortality and 3.27 for circulatory mortality [16]. Potential explanations for the observed differences are the use of very selected reference groups, such as people with a score of 0 on the Centre for Epidemiologic Studies of Depression scale [7, 14] and differences in the study populations [16, 19].

Our study uniquely extends these findings to cancer mortality and causes of death other than circulatory disease and cancer. In patients with diabetes, a previous study reported an increased risk of non-CVD, non-cancer mortality in people with comorbid depression and diabetes, whereas there was no association with CVD and cancer mortality [21]. Since this study was based on patients with diabetes, the joint effect of depression and diabetes could not be examined.

### Possible explanations for our findings

The underlying mechanisms for the synergistic effect of depression and diabetes on mortality risk remain to be established. Since we found synergistic effects of depression and diabetes for different causes of mortality, it is unlikely that the underlying mechanism of the synergistic effects of depression and diabetes is organ-or disease-specific [29]. A more general explanation for the excess mortality among those affected by both depression and diabetes is that depression might make adoption and maintenance of healthy lifestyles, including smoking cessation and self-management more difficult. For example, depression has been shown to be a risk factor for medical noncompliance among individuals with comorbidities [5, 6] which might lead to adverse effects such as poor glycaemic control among individuals with comorbid depression and diabetes [4]. Second, individuals with mental-physical comorbidity may receive sub-optimal quality of care which in turn might increase their risk of adverse events [30, 31]. As such, the negative consequences of depression and diabetes may be aggravated among those with comorbid depression and diabetes due to the lack of successful treatment or self-management strategies for both conditions. However, more research is needed to further explore this hypothesis.

### Implications of this study and future research

Our findings highlight scope for improved care and treatment of depression, particularly in those with diabetes. Despite guidelines that encourage psychological well-being in people with diabetes [33], depression continues to be overlooked in clinical practice [34]. Screening for depression in clinical practice, particularly among those with comorbid depression and diabetes, may be a helpful first step to identify patients at high risk of adverse effects. However, one of the most challenging screening criteria to meet will be provision of cost-effective interventions to individuals identified as being at high risk of adverse events [35-37]. A randomized controlled trial has shown that allocating a trained depression care manager and offering an antidepressant and interpersonal psychotherapy to patients with comorbid depression and diabetes may reduce the 5-year mortality rate [38]. However, the statistical methods used by Bogner, Morales, Post and Bruce [38] were criticized, as they may have resulted in model overfitting [39] and few health systems are likely to have the resources to provide such interventions to the large number of people that might be eligible. Thus, further randomized controlled trials are needed to identify cost-effective interventions that reduce risk of mortality and improve quality of life in patients with one or both of depression and diabetes.

A particular focus of future studies should be on the potential synergistic effect of depression and diabetes not only on circulatory mortality but also on cancer mortality and mortality due to other causes, since this is the first study to report on this. It would be helpful to establish whether the synergistic effect of depression and diabetes on mortality risk is observed in other settings and for participants with type 1 and type 2 diabetes. Furthermore, future studies should attempt to identify mechanisms that may be responsible for the synergistic effect of depression and diabetes on risk of mortality in order to inform development and testing of interventions. Lastly, the temporality of depression and diabetes deserves further attention with one recent study suggesting smaller joint effects of depression and diabetes when both disorders are ascertained at the same point in time compared to analyses assessing the joint effect of diabetes and subsequent depressive symptoms on mortality [17].

### Conclusions

In summary, we found that individuals with depression and diabetes were at high risk of all-cause mortality and mortality due to cancer, circulatory disease, and causes other than circulatory disease or cancer. The synergistic effect of depression on all-cause mortality exceeded the expected additive effect due to either disorder alone which was largely driven by cancer and causes other than circulatory disease and cancer. Although some progress has been made in the past, our findings highlight a need for further research and the potential for improved treatment of depression, particularly in people with diabetes.

## Supporting information

ESM

## Data Availability

All bona fide researchers in academic, commercial, and charitable settings can apply to use the UK Biobank resource for health-related research in the public interest (www.ukbiobank.ac.uk/register-apply/).

## ABBREVIATIONS

AP: Attributable proportion due to interaction
RERI: Relative excess risk due to interaction
S: Synergy index

## ACKNOWLEDGEMENTS

This research has been conducted using the UK Biobank Resource under application number 13797.

## Author contributions

RP, CAJ and SW conceived the study, RP performed the data analysis and wrote the draft manuscript, and CAJ and SW reviewed and edited the manuscript. RP is the guarantor of this work and takes responsibility for the contents of the article.

## Conflicts of interest

We declare that we do not have any competing interests.

## Prior presentation

Parts of this study were presented at the Annual Meeting of the European Diabetes Epidemiology Group, Mondorf-Les-Bains, Luxembourg, 11 – 15 May 2019.

## FUNDING

RP is a PhD scholar funded by the University of Edinburgh. Access to the UK Biobank data was funded by a University of Queensland Early Career Researcher grant, awarded to CAJ.

## Notes

### Competing Interest Statement

The authors have declared no competing interest.

### Author Declarations

Analyses of UK Biobank data are conducted under generic approval from the NHS National Research Ethics Service (Ref 11/NW/0382, approval letter dated 17 June 2011).

