## Supplementary material for "Depression, diabetes, their comorbidity and all-cause and cause-specific mortality: a prospective cohort study": ESM

**Electronic supplementary material**

ESM Material S1: Ascertainment of covariates

The following covariates were ascertained through self-report in the touchscreen questionnaire at the baseline assessment. Income was defined as average household income before tax, with participants selecting one of five categories. Education was defined as highest educational attainment, which we categorized into: College or university degree; A levels, O levels, Certificate of Secondary Education or National Vocational Qualification, or equivalent; and none of the above). Area-based deprivation was assessed through the Townsend Deprivation Index (1), which we divided into fifths for this study. Due to small numbers of participants with a non-white ethnicity, we categorised ethnicity as white or other ethnicity. We categorised smoking as never, previous, or current smoker. We defined alcohol intake as safe drinking if men or women consumed ≤ 14 units of alcohol per week and as risky drinking if alcohol intake exceeded 14 units per week (2). In accordance with the International Physical Activity Questionnaire (3), we assigned levels of physical activity as low, moderate or high. We classified oily fish intake as at least once a week, less than once per week, and never. We used information on daily fruit and vegetable intake to define consumption of at least five fruits or vegetables per day (4). Family history of stroke, heart disease, high blood pressure, and severe depression were defined as self-reported illness of mother or father. We calculated body mass index (kg/m$^2$, BMI) based on measured height and weight ascertained at the assessment centre at baseline, and categorised this as: under- or normal weight (<18.5 – 24.9 kg/m$^2$); overweight (25 – 29.9 kg/m$^2$); obese (30 – 34.9 kg/m$^2$); severely obese (35 – 39.9 kg/m$^2$); and morbidly obese (≥40 kg/m$^2$). Since there were so few participants underweight (0.5% of all participants included in the analysis), these were included in the same category as normal weight. We defined hypertension and high cholesterol levels as diagnosis and/or treatment, ascertained through self-report in the touchscreen questionnaire or nurse interview. Additionally, we identified participants with hypertension through blood pressure measurements of ≥140/90 mmHg at the assessment centre at baseline. We ascertained history of CVD (stroke, myocardial infarction, angina or transient ischaemic attack) and history of cancer using responses to the touchscreen questionnaire and nurse interview, and from linked hospital admission records prior to baseline.


ESM Table S1: Hazard ratios (95% CI) for CVD mortality among UK Biobank participants with neither, one of both of depression and diabetes with truncation of follow-up at six years

| | | **Unadjusted HR (95%CI)** | **Adjusted HR (95%CI) Model 1*** | **Adjusted HR (95%CI) Model 2†** |
|---|---|---|---|---|
| **CVD mortality** | Neither depression, nor diabetes | 1.0 | 1.0 | 1.0 |
| | Depression alone | 1.41 (1.14 – 1.74) | 1.51 (1.22 – 1.87) | 1.24 (1.00 – 1.54) |
| | Diabetes alone | 4.94 (4.19 – 5.83) | 2.74 (2.31 – 3.24) | 1.95 (1.62 – 2.34) |
| | Depression and diabetes | 9.39 (6.97 – 12.64) | 5.25 (3.88 – 7.09) | 3.00 (2.19 – 4.11) |

*Model 1: Age, sex, ethnicity, education, income, area-based deprivation

†Model 2: Model 1 + BMI, physical activity, alcohol intake, smoking, fruit and vegetable intake, oily fish intake, cholesterol, hypertension, cardiovascular disease, cancer, family history of cardiovascular disease, and family history of depression

ESM Table S2: Baseline characteristics and causes of death, separately for UK Biobank participants with and without complete data available

| | **Incomplete cases** (n = 155,761) | **Complete cases** (n = 344,069) |
|---|---|---|
| **Men**, n (%) | 57,446 (36.9) | 170,348 (49.5) |
| **Age** (median [IQR]) | 59.0 [51.0, 64.0] | 57.0 [50.0, 63.0] |
| **White ethnicity**, n (%) | 141,511 (90.9) | 328771 (95.6) |
| **Income in £**, n (%) | | |
| Greater than 100,000 | 1,766 (2.2) | 21,119 (6.1) |
| 52,000 to 100,000 | 10,284 (13.0) | 75,791 (22.0) |
| 31,000 to 51,999 | 18,622 (23.5) | 91,852 (26.7) |
| 18,000 to 30,999 | 22,108 (27.9) | 85,635 (24.9) |
| Less than 18,000 | 26,364 (33.3) | 69,672 (20.2) |
| **Highest educational attainment**, n (%) | | |
| College or university degree | 32,666 (22.4) | 127,578 (37.1) |
| Other degrees* | 74,374 (51.0) | 170,370 (49.5) |
| None of the above | 38,653 (26.5) | 46,121 (13.4) |
| **Area-based deprivation**, n (%) | | |
| 1 = least deprived | 28,679 (18.5) | 71,716 (20.8) |
| 2 | 28,929 (18.6) | 70,868 (20.6) |
| 3 | 29,983 (19.3) | 70,022 (20.4) |
| 4 | 30,356 (19.6) | 69,485 (20.2) |
| 5 = most deprived | 37,196 (24.0) | 61,978 (18.0) |
| **Body mass index**, n (%) | | |
| Under- or normal weight | 47,239 (30.9) | 117,085 (34.0) |
| Overweight | 61,893 (40.5) | 149,255 (43.4) |
| Obese | 29,546 (19.3) | 57,393 (16.7) |
| Severely obese | 9,730 (6.4) | 15,029 (4.4) |
| Morbidly obese | 4,285 (2.8) | 5,307 (1.5) |
| **Physical activity**, n (%) | | |
| High | 45,348 (34.0) | 129,581 (37.7) |
| Moderate | 52,871 (39.6) | 141,478 (41.1) |
| Low | 35,241 (26.4) | 73,010 (21.2) |
| **Alcohol intake** = Risky drinking, n (%) | 29,266 (38.6) | 170,009 (49.4) |
| **Smoking status**, n (%) | | |
| Never | 87,498 (57.2) | 184,918 (53.7) |
| Previous | 47,500 (31.1) | 124,732 (36.3) |
| Current | 17,847 (11.7) | 34,419 (10.0) |
| **Fruit and vegetable intake per day** = Less than five a day, n (%) | 104,873 (68.1) | 241,968 (70.3) |
| **Oily fish intake**, n (%) | | |
| At least once a week | 82,387 (54.3) | 194,921 (56.7) |
| Less than once a week | 49,238 (32.4) | 114,824 (33.4) |
| Never | 20,115 (13.3) | 34,324 (10.0) |
| **History of CVD**, n (%) | 13,067 (8.4) | 22,470 (6.5) |
| **History of cancer**, n (%) | 14,498 (9.3) | 29,828 (8.7) |
| **Hypertension**, n (%) | 91,238 (58.6) | 191,015 (55.5) |
| **High cholesterol levels**, n (%) | 32,544 (20.9) | 62,926 (18.3) |
| **Family history of CVD**, n (%) | 106,112 (68.1) | 239,848 (69.7) |
| **Family history of depression**, n (%) | 12,656 (8.1) | 31,425 (9.1) |
| **Cause of death**, n (%) | | |
| Cancer | 2,872 (1.8) | 5,104 (1.5) |
| Circulatory disease | 1,079 (0.7) | 1,748 (0.5) |
| Other causes | 1,212 (0.8) | 1,709 (0.5) |

* A levels, O levels, Certificate of secondary education, National vocational qualification, or equivalent

ESM Table S3: Hazard ratios (95% CI) for all-cause and cause-specific mortality among UK Biobank participants with neither, one of both of depression and diabetes (complete case analysis)

| | | **Unadjusted HR (95% CI)** | **Adjusted HR (95% CI) Model 1*** | **Adjusted HR (95% CI) Model 2†** |
|---|---|---|---|---|
| **All-cause mortality** | Neither depression, nor diabetes | 1.0 | 1.0 | 1.0 |
| | Depression alone | 1.36 (1.26 – 1.46) | 1.37 (1.27 – 1.47) | 1.19 (1.11 – 1.29) |
| | Diabetes alone | 2.81 (2.61 – 3.02) | 1.79 (1.67 – 1.93) | 1.56 (1.45 – 1.69) |
| | Depression and diabetes | 4.69 (4.07 – 5.41) | 3.02 (2.62 – 3.48) | 2.32 (2.00 – 2.69) |
| **Cancer mortality** | Neither depression, nor diabetes | 1.0 | 1.0 | 1.0 |
| | Depression alone | 1.05 (0.94 – 1.16) | 1.05 (0.94 – 1.16) | 0.93 (0.83 – 1.03) |
| | Diabetes alone | 2.00 (1.80 – 2.22) | 1.36 (1.23 – 1.52) | 1.28 (1.14 – 1.43) |
| | Depression and diabetes | 2.82 (2.24 – 3.54) | 1.96 (1.56 – 2.47) | 1.70 (1.35 – 2.15) |
| **Circulatory mortality** | Neither depression, nor diabetes | 1.0 | 1.0 | 1.0 |
| | Depression alone | 1.37 (1.16 – 1.62) | 1.47 (1.24 – 1.74) | 1.22 (1.02 – 1.44) |
| | Diabetes alone | 4.71 (4.13 – 5.38) | 2.61 (2.28 – 2.99) | 1.82 (1.57 – 2.10) |
| | Depression and diabetes | 7.55 (5.83 – 9.79) | 4.31 (3.32 – 5.59) | 2.41 (1.84 – 3.16) |
| **Mortality due to other causes** | Neither depression, nor diabetes | 1.0 | 1.0 | 1.0 |
| | Depression alone | 2.47 (2.16 – 2.82) | 2.37 (2.07 – 2.72) | 2.11 (1.84 – 2.43) |
| | Diabetes alone | 3.68 (3.16 – 4.28) | 2.24 (1.92 – 2.61) | 2.10 (1.77 – 2.48) |
| | Depression and diabetes | 8.36 (6.47 – 10.79) | 4.83 (3.73 – 6.25) | 3.85 (2.93 – 5.06) |

*Model 1: Age, sex, ethnicity, education, income, area-based deprivation

†Model 2: Model 1 + BMI, physical activity, alcohol intake, smoking, fruit and vegetable intake, oily fish intake, cholesterol, hypertension, cardiovascular disease, cancer, family history of cardiovascular disease, and family history of depression

ESM Table S4: Hazard ratios (95% CI) for all-cause and cause-specific mortality among UK Biobank participants with neither, one of both of depression and diabetes, separately for men and women (complete case analysis)

| | | **Men** Adjusted HR (95% CI)* | **Women** Adjusted HR (95% CI)* |
|---|---|---|---|
| **All-cause mortality** | Neither depression, nor diabetes | 1.0 | 1.0 |
| | Depression alone | 1.24 (1.12 – 1.38) | 1.15 (1.03 – 1.29) |
| | Diabetes alone | 1.58 (1.44 – 1.72) | 1.53 (1.28 – 1.83) |
| | Depression and diabetes | 2.40 (2.02 – 2.84) | 2.09 (1.56 – 2.81) |
| **Cancer mortality** | Neither depression, nor diabetes | 1.0 | 1.0 |
| | Depression alone | 0.87 (0.73 – 1.02) | 0.99 (0.86 – 1.14) |
| | Diabetes alone | 1.28 (1.12 – 1.46) | 1.27 (1.01 – 1.60) |
| | Depression and diabetes | 1.86 (1.41 – 2.45) | 1.38 (0.88 – 2.17) |
| **Circulatory mortality** | Neither depression, nor diabetes | 1.0 | 1.0 |
| | Depression alone | 1.30 (1.06 – 1.59) | 1.03 (0.74 – 1.43) |
| | Diabetes alone | 1.75 (1.50 – 2.06) | 2.35 (1.62 – 3.40) |
| | Depression and diabetes | 2.53 (1.88 – 3.40) | 1.95 (0.97 – 3.92) |
| **Mortality due to other causes** | Neither depression, nor diabetes | 1.0 | 1.0 |
| | Depression alone | 2.16 (1.81 – 2.58) | 2.01 (1.61 – 2.52) |
| | Diabetes alone | 2.18 (1.81 – 2.63) | 1.84 (1.22 – 2.76) |
| | Depression and diabetes | 3.49 (2.51 – 4.84) | 4.76 (2.91 – 7.80) |

*Fully adjusted model: Age, ethnicity, education, income, area-based deprivation, BMI, physical activity, alcohol intake, smoking, fruit and vegetable intake, oily fish intake, cholesterol, hypertension, cardiovascular disease, cancer, family history of cardiovascular disease, and family history of depression

ESM Table S5: Measures of additive and multiplicative interaction between depression and diabetes on risk of all-cause and cause-specific mortality (complete case analysis)

| | **Additive interaction** | | | **Multiplicative interaction** |
|---|---|---|---|---|
| | ***RERI (CI)*** | ***AP (CI)*** | ***S (CI)*** | ***p-value*** |
| **All-cause mortality** | 0.56 (0.21 – 0.92) | 0.24 (0.12 – 0.36) | 1.74 (1.28 – 2.37) | 0.003 |
| **Cancer mortality** | 0.38 (-0.30 – 1.06) | 0.16 (-0.09 – 0.40) | 1.37 (0.81 – 2.31) | 0.005 |
| **Circulatory mortality** | 0.50 (0.08 – 0.92) | 1.37 (0.81 – 2.31) | 3.47 (1.27 – 9.49) | 0.294 |
| **Mortality due to other causes** | 0.64 (-0.41 – 1.70) | 0.17 (-0.07 – 0.40) | 1.29 (0.88 – 1.90) | 0.658 |
| **CVD mortality** | 0.45 (-0.38 – 1.28) | 0.17 (-0.10 – 0.44) | 1.38 (0.80 – 2.36) | 0.321 |
| **Non-CVD circulatory mortality** | 0.17 (-0.99 – 1.32) | 0.10 (-0.52 – 0.72) | 1.30 (0.23 – 7.41) | 0.697 |

AP: Attributable proportion, RERI: Relative excess risk for interaction, S: Synergy index

ESM Table S6: Hazard ratios (95% CI) for CVD- and non-CVD circulatory mortality among UK Biobank participants with neither, one of both of depression and diabetes (complete case analysis)

| | | **Unadjusted HR (95%CI)** | **Adjusted HR (95%CI) Model 1*** | **Adjusted HR (95%CI) Model 2†** |
|---|---|---|---|---|
| **CVD mortality** | Neither depression, nor diabetes | 1.0 | 1.0 | 1.0 |
| | Depression alone | 1.37 (1.13 – 1.67) | 1.47 (1.21 – 1.79) | 1.20 (0.98 – 1.46) |
| | Diabetes alone | 5.25 (4.53 – 6.08) | 2.87 (2.47 – 3.34) | 2.00 (1.70 – 2.36) |
| | Depression and diabetes | 8.43 (6.32 –11.23) | 4.69 (3.51 – 6.27) | 2.65 (1.96 – 3.59) |
| **Non-CVD circulatory mortality** | Neither depression, nor diabetes | 1.0 | 1.0 | 1.0 |
| | Depression alone | 1.36 (0.98 – 1.88) | 1.48 (1.07 – 2.06) | 1.27 (0.91 – 1.78) |
| | Diabetes alone | 3.24 (2.40 – 4.37) | 1.87 (1.38 – 2.54) | 1.27 (0.92 – 1.77) |
| | Depression and diabetes | 5.17 (2.84 – 9.43) | 3.16 (1.73 – 5.79) | 1.71 (0.92 – 3.20) |

ESM Table S7: Causes of death in the other mortality group, overall and separately for participants with none, one or both of depression and diabetes (n, %)

| **Causes of death** | **Neither depression, nor diabetes** (n = 1,962) | **Depression alone** (n = 468) | **Diabetes alone** (n = 380) | **Depression and diabetes** (n = 111) | **Total** (n = 2,921) |
|---|---|---|---|---|---|
| Diseases of the respiratory system | 517 (26.4%) | 132 (28.2%) | 101 (26.6%) | 24 (21.6%) | 774 (26.5%) |
| Diseases of the digestive system | 343 (17.5%) | 88 (18.8%) | 76 (20.0%) | 23 (20.7%) | 530 (18.1%) |
| External causes of morbidity and mortality | 320 (16.3%) | 107 (22.9%) | 37 (9.7%) | 10 (9.0%) | 474 (16.2%) |
| Diseases of the nervous system | 325 (16.6%) | 59 (12.6%) | 31 (8.2%) | 11 (9.9%) | 426 (14.6%) |
| Certain infectious and parasitic diseases | 90 (4.6%) | 12 (2.6%) | 14 (3.7%) | 3 (2.7%) | 119 (4.1%) |
| Endocrine, nutritional and metabolic diseases | 36 (1.8%) | 7 (1.5%) | 58 (15.3%) | 18 (16.2%) | 119 (4.1%) |
| Mental and behavioural disorders | 69 (3.5%) | 16 (3.4%) | 9 (2.4%) | 1 (0.9%) | 95 (3.3%) |
| Neoplasms (in situ, benign, uncertain or unknown behaviour) | 64 (3.3%) | 9 (1.9%) | 9 (2.4%) | 3 (2.7%) | 85 (2.9%) |
| Diseases of the musculoskeletal system and connective tissue | 57 (2.9%) | 13 (2.8%) | 7 (1.8%) | 5 (4.5%) | 82 (2.8%) |
| Diseases of the genitourinary system | 37 (1.9%) | 7 (1.5%) | 21 (5.5%) | 6 (5.4%) | 71 (2.4%) |
| Symptoms, signs and abnormal clinical and laboratory findings, not elsewhere classified | 43 (2.2%) | 8 (1.7%) | 10 (2.6%) | 4 (3.6%) | 65 (2.2%) |
| Diseases of the blood and blood-forming organs and certain disorders involving the immune mechanism | 20 (1.0%) | 2 (0.4%) | 5 (1.3%) | 1 (0.9%) | 28 (1.0%) |
| Congenital malformations, deformations and chromosomal abnormalities | 22 (1.1%) | 4 (0.9%) | 1 (0.3%) | 0 (0.0%) | 27 (0.9%) |
| Other death (temporary code) | 12 (0.6%) | 1 (0.2%) | 0 (0.0%) | 1 (0.9%) | 14 (0.5%) |
| Diseases of the skin and subcutaneous tissue | 7 (0.4%) | 3 (0.6%) | 1 (0.3%) | 1 (0.9%) | 12 (0.4%) |

ESM Table S8: Measures of additive and multiplicative interaction between depression and diabetes on risk of all-cause and cause-specific mortality

| | **Additive interaction** | | | **Multiplicative interaction** |
|---|---|---|---|---|
| | ***RERI (CI)*** | ***AP (CI)*** | ***S (CI)*** | ***p-value*** |
| **All-cause mortality** | 0.29 (0.03 – 0.54) | 0.13 (0.03 – 0.24) | 1.32 (1.05 – 1.67) | 0.182 |
| **Cancer mortality** | 0.38 (0.07 – 0.68) | 0.23 (0.08 – 0.39) | 2.57 (1.27 – 5.23) | 0.006 |
| **Circulatory mortality** | -0.03 (-0.52 – 0.46) | -0.01 (-0.24 – 0.21) | 0.98 (0.65 – 1.46) | 0.578 |
| **Mortality due to other causes** | 0.26 (-0.48 – 1.01) | 0.07 (-0.12 – 0.27) | 1.11 (0.83 – 1.49) | 0.061 |
| **CVD mortality** | -0.26 (-0.85 – 0.33) | -0.11 (-0.39 – 0.17) | 0.83 (0.54 – 1.29) | 0.197 |
| **Non-CVD circulatory mortality** | 0.57 (-0.31 – 1.46) | 0.29 (-0.06 – 0.63) | 2.33 (0.68 – 7.98) | 0.177 |

AP: Attributable proportion, RERI: Relative excess risk for interaction, S: Synergy index

ESM Table S9: Hazard ratios (95% CI) for CVD- and non-CVD circulatory mortality among UK Biobank participants with neither, one of both of depression and diabetes

| | | **Unadjusted HR (95%CI)** | **Adjusted HR (95%CI) Model 1*** | **Adjusted HR (95%CI) Model 2†** |
|---|---|---|---|---|
| **CVD mortality** | Neither depression, nor diabetes | 1.0 | 1.0 | 1.0 |
| | Depression alone | 1.52 (1.32 – 1.76) | 1.62 (1.39 – 1.88) | 1.30 (1.12 – 1.52) |
| | Diabetes alone | 5.82 (5.21 – 6.49) | 3.27 (2.92 – 3.66) | 2.25 (1.99 – 2.55) |
| | Depression and diabetes | 6.96 (5.54 – 8.75) | 4.20 (3.33 – 5.29) | 2.29 (1.80 – 2.92) |
| **Non-CVD circulatory mortality** | Neither depression, nor diabetes | 1.0 | 1.0 | 1.0 |
| | Depression alone | 1.33 (1.04 – 1.71) | 1.41 (1.09 – 1.81) | 1.19 (0.92 – 1.54) |
| | Diabetes alone | 3.07 (2.45 – 3.86) | 1.88 (1.49 – 2.37) | 1.24 (0.96 – 1.59) |
| | Depression and diabetes | 5.89 (3.97 – 8.74) | 3.92 (2.63 – 5.83) | 2.00 (1.31 – 3.06) |

*Model 1: Age, sex, ethnicity, education, income, area-based deprivation
†Model 2: Model 1 + BMI, physical activity, alcohol intake, smoking, fruit and vegetable intake, oily fish intake, cholesterol, hypertension, cardiovascular disease, cancer, family history of cardiovascular disease, and family history of depression

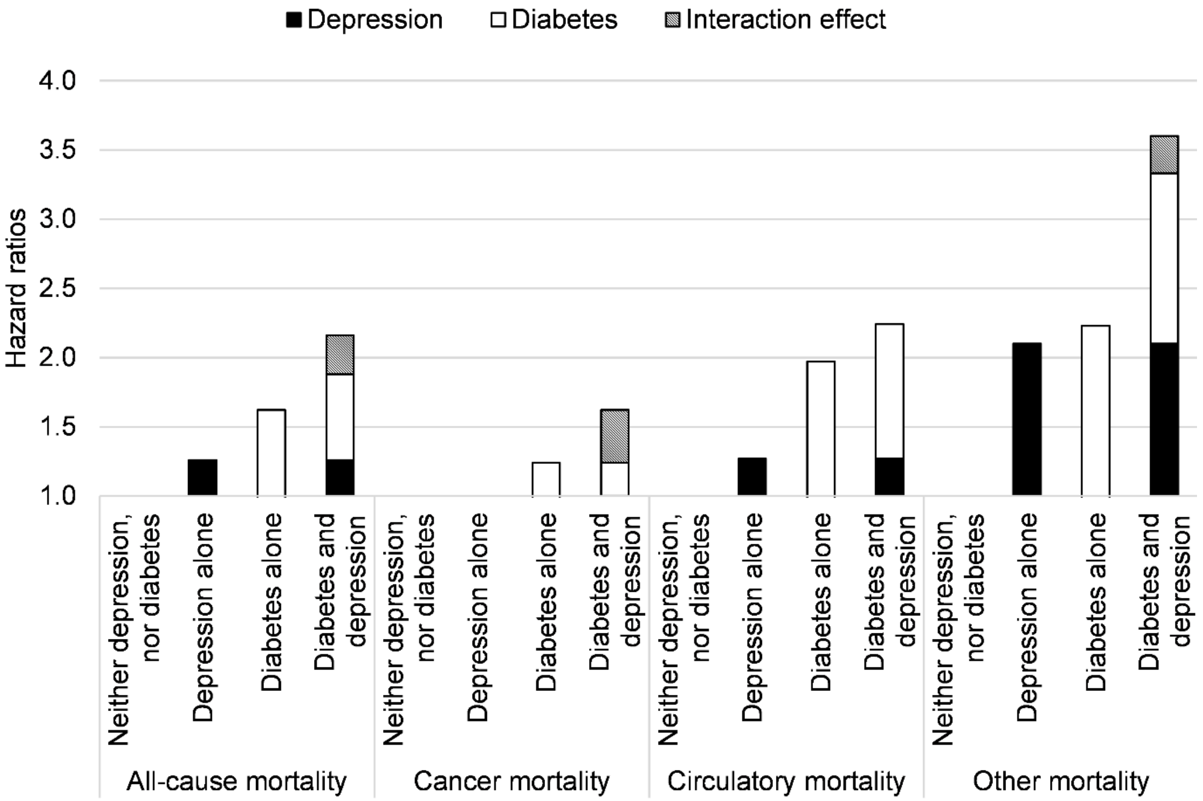


ESM Fig. S1: Hazard ratios (95% CI) for all-cause and cause-specific mortality among UK Biobank participants with neither, one of both of depression and diabetes, separately for men and women (fully adjusted models*)

*Fully adjusted model: Age, ethnicity, education, income, area-based deprivation, BMI, physical activity, alcohol intake, smoking, fruit and vegetable intake, oily fish intake, cholesterol, hypertension, cardiovascular disease, cancer, family history of cardiovascular disease, and family history of depression
